# Pre-exposure prophylaxis adherence with real-time adherence feedback and partner HIV self-testing: A pilot trial among postpartum women

**DOI:** 10.1101/2021.07.02.21259896

**Authors:** Dvora Leah Joseph Davey, Kathryn Dovel, Rufaro Mvududu, Dorothy Nyemba, Nyiko Mashele, Linda-Gail Bekker, Pamina M. Gorbach, Thomas J. Coates, Landon Myer

## Abstract

**Background:** Pre-exposure prophylaxis (PrEP) is safe and effective in postpartum women. HIV self-testing (HIVST) for partners combined with biofeedback counselling through real-time adherence measures may improve daily PrEP use among postpartum women.

**Methods:** Between August 2020 and April 2021 we conducted a pilot study in one primary care clinic in Cape Town, South Africa. We randomized postpartum women who initiated PrEP in pregnancy 1:1 to the intervention group (HIVST + biofeedback counselling following urine tenofovir test) or to standard of care (facility-based HIV tests and routine counselling without biofeedback). The outcomes of interest were PrEP adherence in the past 48-72hours via urine tenofovir tests and partner HIV testing, measured 1-month after randomisation. Secondary outcomes included proportion of partners who tested for HIV and discrepancy between self-reported PrEP adherence and urine tenofovir result.

**Findings:** We enrolled 106 women (median age=26 years; median months postpartum=2). Almost half of women reported having sex since giving birth (48%); 76% of those reported condomless sex at last sex. At enrolment most women (72%) reported missing <2 doses in the past 7-days; 36% of women had tenofovir present in her urine (no significant differences by arm). One month after enrolment, 62% (n=33) of women in the intervention arm had tenofovir present in their urine compared to 34% (n=18) in the standard of care arm (RR=1.83; 95% CI=1.19, 2.82). Two-thirds of women in the intervention arm reported that her partner tested for HIV (66%; n=35); compared to 17% (n=9) in the standard of care arm (RR=3.89; 95% CI=2.08, 7.27). The proportion of women with a discrepant adherence result (self-reported good recent adherence with no tenofovir in urine test) was significantly lower in the intervention group (n=8; 17%) compared to the standard of care group (n=24; 46%) (RR=0.33; 95% CI=0.17, 0.67). No social or clinical adverse events were reported in the intervention arm.

**Interpretation:** In this pilot study, HIVST for partners and biofeedback counseling increased levels of recent PrEP adherence, pointing to the importance of these interventions to support PrEP use in this population.

**Funding:** Our study is funded by Fogarty International Center (K01TW011187) with additional support from NIMH (R01MH116771). Trial registration: Clinicaltrials.gov (NCT04897737). Funders had no role in data collection or analysis.

## Introduction

Over half of HIV infections globally occur among cisgender women^1^ and the risk of HIV acquisition nearly doubles during periods of pregnancy and postpartum due to biological and behavioral factors.^2-4^ Few primary HIV prevention interventions exist for the majority of pregnant or breastfeeding women (PBFW) who initially test HIV-negative in antenatal care (ANC). With adequate adherence, oral pre-exposure prophylaxis (PrEP) is highly effective for HIV prevention, is safe and feasible for use in PBFW without HIV to prevent HIV acquisition during pregnancy and the postpartum period.^5^ Despite its potential value for HIV prevention, PrEP adherence is low in many settings,^6-10^ including in PBFW ^11^.

Preliminary findings from our study with PBFW in South Africa show high levels of PrEP initiation (>90% of eligible women starting PrEP), but low levels of continuation and adherence on PrEP (<60% of women who initiated PrEP continued and were adherent in the postpartum period).^12^ Given the high HIV incidence among PBFW and risk of infant HIV acquisition, new intervention strategies are urgently needed to improve adherence among PBFW at risk for HIV.^13-15^ Primary barriers to daily oral PrEP among PBFW include low percieved risk of infection and couple dynamics, suboptimal counseling and adherence strategies, and facility-level barriers to PrEP access.^14,16^ Women often have limited knowledge about their sexual partner’s serostatus, and many women underestimate the likelihood that their partner is HIV-infected^12,17^, others report needing permission from their partner to use PrEP and fear disclosing to their partner that they are taking PrEP.^17^ PrEP use in PBFW requires monthly or bi-monthly facility visits with HIV testing throughout the duration of PrEP use, and PrEP consultations are likely the only reason women visit the clinic once postpartum, adding substantial burden.^17^ Further, standard PrEP counselling is based on self-reported adherence, which may over-report true drug adherence,^18-22^ rather than focusing on addressing barriers to daily PrEP use (and promoting condom use when not adherent) to protect the participant.^22^

We piloted a combination intervention aimed to address barriers to PrEP. First, empowering women to monitor their own HIV testing and increase knowledge of their partner’s serostatus using HIV self-testing (HIVST) can alleviate the need for multiple clinical visits whilst improving HIV risk perception, disclosure, and partner support for daily PrEP taking. Second, PrEP adherence counselling based on recent real-time adherence levels may improve adherence to daily PrEP. A recently developed immunoassay using urine measures tenofovir (TFV) and is sensitive (96%) and specific (100%) when compared to plasma levels. ^22-26^ The urine assay shows TFV concentrations if PrEP is taken in the past 48-72 hours and is processed within 10-15 minutes, enabling providers to adapt counselling messages immediately,^22,26^ potentially increasing motivation for adherence. We conducted a randomized control pilot trial to test the impact of a combined intervention (HIVST for PrEP users and partners, and real-time adherence biofeedback) on recent PrEP adherence among postpartum women who took PrEP during pregnancy compared to standard of care (SOC).

## Methods

This pilot study (Clinical Trial NCT04897737) was designed as a parallel-arm randomized control trial, embedded within an ongoing parent study, PrEP-PP (PrEP in Pregnant and Postpartum women). PrEP-PP is an open prospective cohort which enrols consenting pregnant, HIV-uninfected adolescent girls and women (age ≥16 years) at the first ANC visit and follows participants through 12-months post-delivery at a single public health clinic in Cape Town, South Africa^12^.

### Study procedures

Between August 2020 and April 2021, trained study staff recruited women from the PrEP-PP cohort for the pilot study. Eligibility criteria included having given birth to a live infant in the preceding 4-24 weeks, documented HIV-negative status in the study on the date of screening,^27^ initiating PrEP in the recently completed pregnancy, and reported having at least one sexual partner. Women were randomized in 1:1 to the combination intervention arm (n=53) or SOC arm (n=53) through a series of opaque envelopes kept secure by the study coordinator; random assignments were generated by the PI using a computer programme.

### Standard of care

Women received the SOC national PrEP services including HIV counselling and testing at the facility by a trained HIV counsellor, face-to-face counselling that is based on self-reported adherence, and HIV testing referral slips to invite sex partners for HIV testing. Per national guidelines, women were expected to return to the facility every 3-months for HIV testing and PrEP dispensing. Women gave a urine sample for the tenofovir test to evaluate recent PrEP adherence, but without feedback on the test result, nor was additional counselling provided in the SOC group, though both groups were told that their urine was being tested for PrEP adherence monitoring.

### Intervention

Based on the Ickovics’ and Meisler’s conceptual framework for clinical care^28^ and formative work^12^, we hypothesized that a combination intervention with HIVST for PrEP users and partners, combined with biofeedback counselling through real-time adherence measures, would improve recent PrEP adherence as compared to SOC (**Supplemental Figure 1**). The intervention package included provision of OraQuick HIVST for the woman and one test for each of her reported sex partners, along with instructions on how to use and interpret the results. Counselors encouraged participants to text a photo of the result of her test with her partner’s test. If the result was reactive, participants were referred for confirmatory HIV testing and linkage to HIV care (and immediate PrEP cessation if the participant’s test was reactive). Prior to PrEP adherence counseling, study counsellors requested that women provide a urine sample that was tested using the UrSure/Orasure^22,26^ test that provides feedback on the result in 10-15 minutes (control line present=at least one dose in previous 48 hours; control and test lines present=no dose in at least previous 48 hours). Following the results, trained counsellors provided biofeedback adherence counselling based on results from the urine lateral flow assay. If tenofovir was detected, women were encouraged to continue to take daily PrEP and discussed what barriers they may face in the coming month to daily adherence, so they can come up with an adherence plan. If tenofovir was undetected, counsellors discussed how quickly tenofovir can pass through the body and that they are not judging the participant’s PrEP use, but want to come up with a plan to improve daily adherence. Women without tenofovir were encouraged to use condoms at next sexual encounter, and to take PrEP for 7-days prior to condomless sex. Counsellors asked women to report partner HIVST result within 1-month after distribution by sending a picture of the HIVST test result or bringing the used HIVST kit to study staff to confirm that he had tested.

### Follow-up

Both arms were asked to return in 1-month to have additional HIV testing, adherence counselling and urine tenofovir testing for PrEP. If they did not return, they were offered a phone interview for the questionnaire (and urine tenofovir testing for PrEP was missing).

### Measures

Questionnaires were collected at baseline and follow up visit for all participants included items on: (a) demographic information, (b) partner HIV testing, (c) sexual behaviours, (d) intimate partner violence (using the WHO IPV scale^29^), (e) PrEP adherence according to self-report (seven- and 30-day recall), and (f) the acceptability of the intervention (in the intervention group) using a Likert scale stratified by their intervention outcome. We collected data on verification of use of HIVST though a cell phone photograph or bringing the test back at the following visit.

The primary outcome was recent PrEP adherence measured though point of care tenofovir detection in urine (reflecting adherence in past 48-72 hours). Secondary outcomes included: (a) proportion of sexual partners who tested for HIV within the 1-month study follow-up period based on reporting by the participant, confirmed with SMS/WhatsApp picture or return of used HIVST for intervention arm participants or by self-report from participant in the control arm, and (b) discrepancy between self-reported PrEP adherence urine tenofovir measure of recent PrEP adherence.

### Data analysis

We examined baseline demographic and behavioural characteristics by study arm using medians and interquartile ranges (IQR) for continuous variables, and frequencies and proportions for categorical variables. Wilcoxon rank-sum (for continuous variables), Chi-squared, and Fischer’s exact tests (for categorical variables) were used to explore the characteristics by study arm. We also analysed differences in participants retained versus lost to follow-up. All statistical tests were two-sided at α=0.05.

All analyses were by intention-to-treat. In the case of missing outcomes for participants who did not return for the follow-up visit, we assigned outcome values of: poor PrEP adherence (TFV undetected) and partner not tested for HIV. We constructed univariate Poisson regression models with robust standard errors to examine the predictors of outcomes of interests. Model results are presented as crude risk ratios (RR) and risk differences (RD) with 95% confidence intervals (CIs). All statistical analyses were conducted with STATA v.15 (College Station, TX: StataCorp LLC).

### Ethics

The study was approved by the University of Cape Town Human Research Ethics Committee and the University of California Los Angeles Institutional Review Board. Written informed consent was attained by all participants.

## Results

We screened 176 postpartum women who were enrolled in the PrEP-PP study; 38 (22%) refused participation while 32 (18%) were ineligible (most commonly because they did not report a partner or partner was out of town). We enrolled 106 postpartum women (60% of women screened) in the pilot study and randomized 1:1 to intervention and SOC arm. Overall, 48 of 53 women randomised to the intervention group, and 52 of 53 women randomised to the SOC group, returned for follow-up (91% and 98% respectively) (**Figure 1)**.

**Figure 1.**
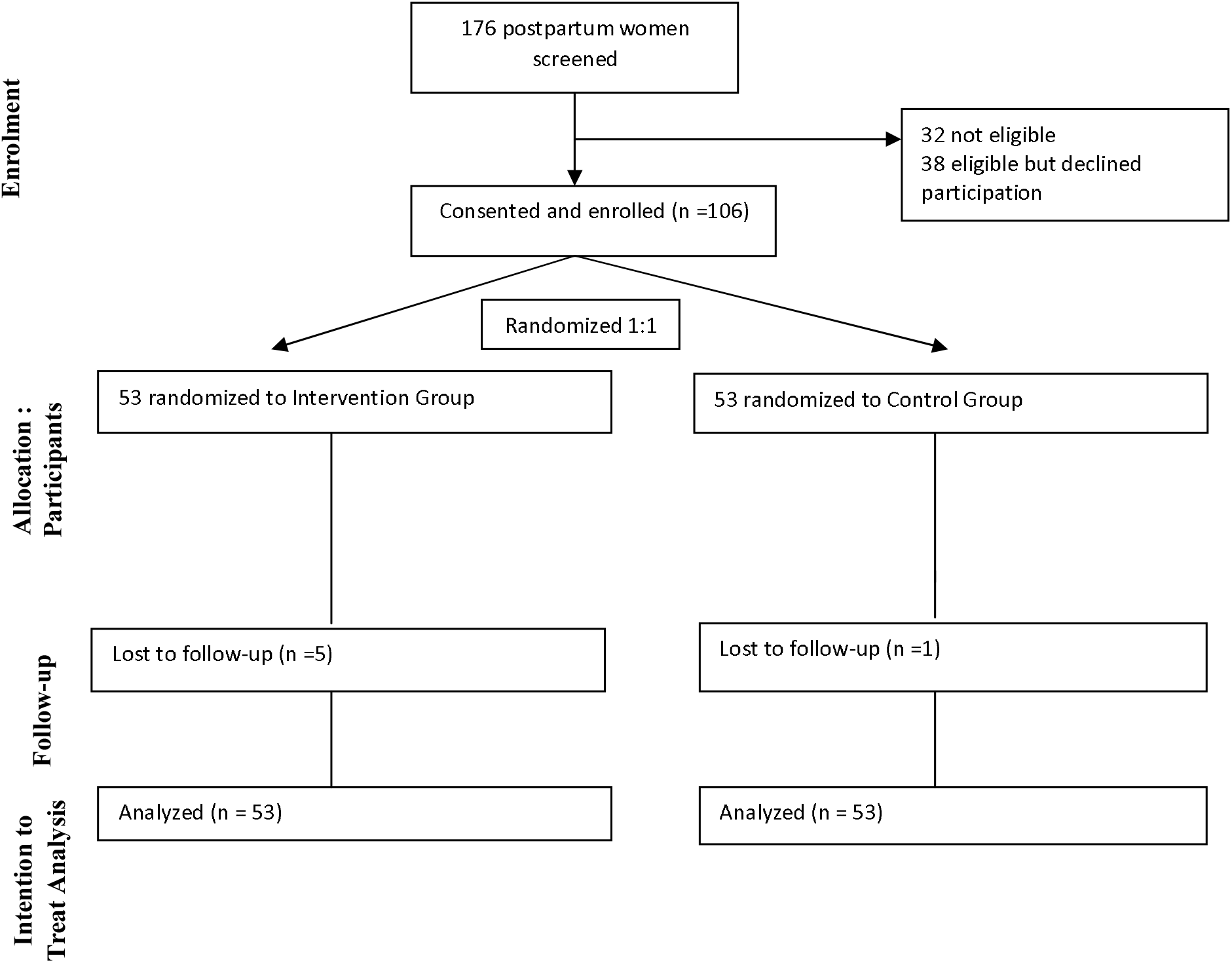
Consort diagram. Participant flow of women screened, enrolled and randomized in the postpartum adherence study

The median age was 26 years (IQR=23-31 years) and women were 2 median months’ postpartum (IQR=1-6-months). Half of women had some secondary school education (n=56; 53%) and 75% were unemployed. Most women were unmarried or non-cohabitating with their partner (56%). Overall, 85% reported they knew their partner’s serostatus. Almost half of women reported having sex since giving birth (48%), and 76% of those reported having condomless sex at last sex. Overall, 72% of women reported missing 0-1 PrEP doses in the past week, and 28% reported missing 2 or more doses. There was no difference in self-reported PrEP adherence at baseline by arm.

Using urine assays as a biomarker for baseline tenofovir levels, 36% (n=68) of women had tenofovir present in their urine. Overall, almost half (43%; n=46) had discrepant results, meaning that they reported good recent adherence (missing <2 doses in the past week) but did not have tenofovir in her urine for the same timeframe (**Table 1**).

**Table 1.** Baseline demographic and behavioural factors by study arm in Cape Town postpartum women (N=106 women), November 2020-April 2021

|  | **Total**<br>**(N=106)** | **Total**<br>**(%)** | **Intervention**<br>**(n=53)** | **Intervention**<br>**(%)** | **Control**<br>**(n=53)** | **Control**<br>**(%)** | **p-value** |
| --- | --- | --- | --- | --- | --- | --- | --- |
| Age, years (median, IQR) | 26 (23-31) |  | 26 (23-31) |  | 26 (23-31) |  | 0.91 |
| Postpartum age, months (median, IQR) | 2 (1-6) |  | 2 (1-6) |  | 2 (1-6) |  | 0.81 |
| Weeks since study enrolment and 1st PrEP prescription (median, IQR) | 25 (13 - 47) |  | 26 (23 - 57) |  | 24 (13 -37) |  | 0.17 |
| Education |  |  |  |  |  |  |  |
| Below secondary | 56 | 53% | 26 | 49% | 30 | 57% | 0.44 |
| Secondary or above | 50 | 47% | 27 | 51% | 23 | 43% |  |
| Employment status |  |  |  |  |  |  |  |
| Full-time | 18 | 17% | 7 | 13% | 11 | 21% | 0.66 |
| Part-time | 7 | 7% | 3 | 6% | 4 | 8% |  |
| Self employed | 1 | 1% | 1 | 2% | 0 | 0% |  |
| Not employed | 79 | 75% | 41 | 79% | 38 | 72% |  |
| Prior pregnancies (#) |  |  |  |  |  |  |  |
| 1 | 38 | 36% | 16 | 30% | 22 | 42% | 0.46 |
| 2--3 | 56 | 53% | 30 | 57% | 26 | 49% |  |
| 4+ | 12 | 11% | 7 | 13% | 5 | 9% |  |
| Relationship status |  |  |  |  |  |  |  |
| No relationship | 1 | 1% | 1 | 2% | 0 | 0% | 0.70 |
| Married | 14 | 13% | 8 | 15% | 6 | 11% |  |
| Cohabiting | 32 | 30% | 14 | 26% | 18 | 34% |  |
| Not cohabiting, not married | 59 | 56% | 30 | 57% | 29 | 55% |  |
| Partner's Education |  |  |  |  |  |  |  |
| Below secondary | 32 | 33% | 14 | 29% | 18 | 37% | 0.39 |
| Secondary or above | 66 | 67% | 35 | 71% | 31 | 63% |  |
| Partner employed |  |  |  |  |  |  |  |
| Full-time | 50 | 47% | 24 | 45% | 26 | 49% | 0.44 |
| Part-time | 26 | 25% | 11 | 21% | 15 | 28% |  |
| Self employed | 2 | 2% | 1 | 2% | 1 | 2% |  |
| Not employed | 27 | 25% | 17 | 32% | 10 | 19% |  |
| Unknown | 1 | 1% | 0 | 0% | 1 | 2% |  |
| Partner HIV status |  |  |  |  |  |  |  |
| HIV negative | 89 | 84% | 46 | 87% | 43 | 81% | 0.42 |
| HIV positive | 1 | 1% | 1 | 2% | 0 | 0% |  |
| Unknown | 16 | 15% | 6 | 11% | 10 | 19% |  |
| Condom use |  |  |  |  |  |  |  |
| Condomless sex at last sex | 81 | 76% | 40 | 75% | 41 | 77% | 0.82 |
| Condom used at last sex | 25 | 24% | 13 | 25% | 12 | 23% |  |
| Number of sex partners in past year |  |  |  |  |  |  |  |
| 0-1 partner | 99 | 93% | 49 | 92% | 50 | 94% | 1.00 |
| >1 partner | 7 | 7% | 4 | 8% | 3 | 6% |  |
| # days missed PrEP in last 7 days |  |  |  |  |  |  |  |
| 0-1 | 76 | 72% | 40 | 75% | 36 | 68% | 0.46 |
| 2--3 | 10 | 9% | 3 | 6% | 7 | 13% |  |
| 3--4 | 20 | 19% | 10 | 19% | 10 | 19% |  |
| # days missed PrEP in last 30 days |  |  |  |  |  |  |  |
| 0-3 | 64 | 60% | 36 | 68% | 28 | 53% | 0.41 |
| 4--7 | 17 | 16% | 7 | 13% | 10 | 19% |  |
| 8--11 | 4 | 4% | 1 | 2% | 3 | 6% |  |
| >11 | 21 | 20% | 9 | 17% | 12 | 23% |  |
| Tenofovir present in urine Tfv test |  |  |  |  |  |  |  |
| Tenofovir absent in urine | 68 | 64% | 34 | 64% | 34 | 64% | 1.00 |
| Tenofovir present in urine | 38 | 36% | 19 | 36% | 19 | 36% |  |

Women who did not return for follow up (n=6) tended to be older and none had tenofovir present in their urine at baseline compared to those retained (p<0.05; **Supplemental Table 1**).

### Outcomes

Median follow up was 3-weeks (IQR=3-7 weeks). Overall, 62% (n=33) of the intervention arm and 34% (n=18) of the SOC arm had tenofovir present in their urine using urine assays, with nearly double the relative risk of a positive tenofovir result for the intervention arm (RR=1.83; 95% CI= 1.19, 2.82). The risk difference between the intervention and SOC groups for recent PrEP adherence was 28.3% (95% CI=10.06, 46.55) (**Figure 2, Supplemental Table 2**).

**Figure 2.**
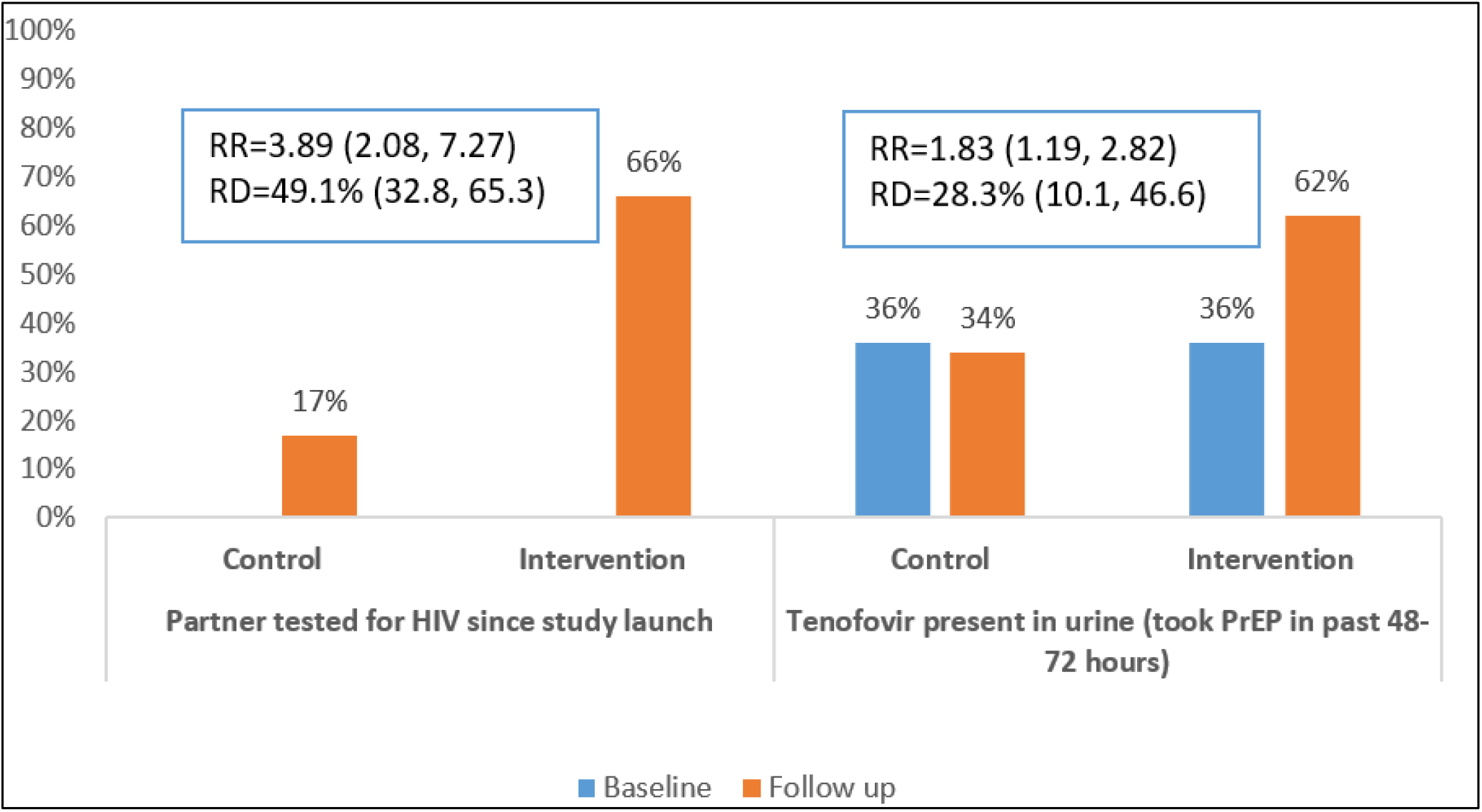
Results from randomized control trial of male partner HIV testing and urine tenofovir testing with self-reported PrEP adherence results in N=106 postpartum women in Cape Town, South Africa

Overall, 66% (n=35) of partners in the intervention and 17% (n=9) of partners in the SOC arm tested for HIV. Women in the intervention group had an almost 4-fold increase of reporting her partner had tested for HIV compared to the SOC group (RR=3.89, 95% CI=2.08, 7.27). The risk difference between women in the intervention vs. control was 49% (95% CI=32.8, 65.3). Among those whose partners tested, most women reported testing with their partner – 83% (n=29 of 35 partners tested) in the intervention arm and 69% (n=6 of 9 tested) in the SOC arm. In each arm one partner tested HIV-positive (8.6% positivity rate for intervention arm, 11% positivity rate for SOC arm).

The proportion of women with a discrepant adherence result (self-reported good recent adherence with no TFV in urine test) was significantly lower in the intervention group (n=8; 17%) compared to the SOC group (n=24; 46%) (RR=0.33, 95% CI=0.17, 0.67). In the intervention group, 77% (n=41) women reported good adherence in the past 7-days (missed <2 doses), vs. 81% in the SOC group (RR=1.06, 95% CI=0.89, 1.26). Postpartum sexual activity was the same in each arm of the study at follow up (RR=0.96, 95% CI=0.56, 1.65) (Not tabled). In addition, 35% of women reported that they greatly improved their PrEP adherence following the intervention.

### Acceptability

At follow-up, 46 women in the intervention arm reported that they used the HIVST to test themselves (96%) and 42 gave a HIVST kit to their partner (87%). Most of those who distributed a HIVST kit (93%) demonstrated how to use the kit to their partner. One woman said she pressured her partner to test; this participant was the only one to report that testing caused conflict in her relationship. The majority of women said that their partner used the HIVST (66% overall and 83% of those who gave their partner the test). Most women reported that the HIVST was not difficult (91%) and that they felt comfortable distributing and demonstrating HIVST kits to their sexual partners. Almost all women whose partner tested (91%) brought in the HIVST or sent in a photo of the test. Of the women who reported that her partner tested, 82% said that they were very satisfied with the partner HIVST kits and counselling on their use. Women whose partners did not test were less satisfied with the HIVST intervention (50% reported that they were very satisfied) (**Table 2**).

**Table 2.**
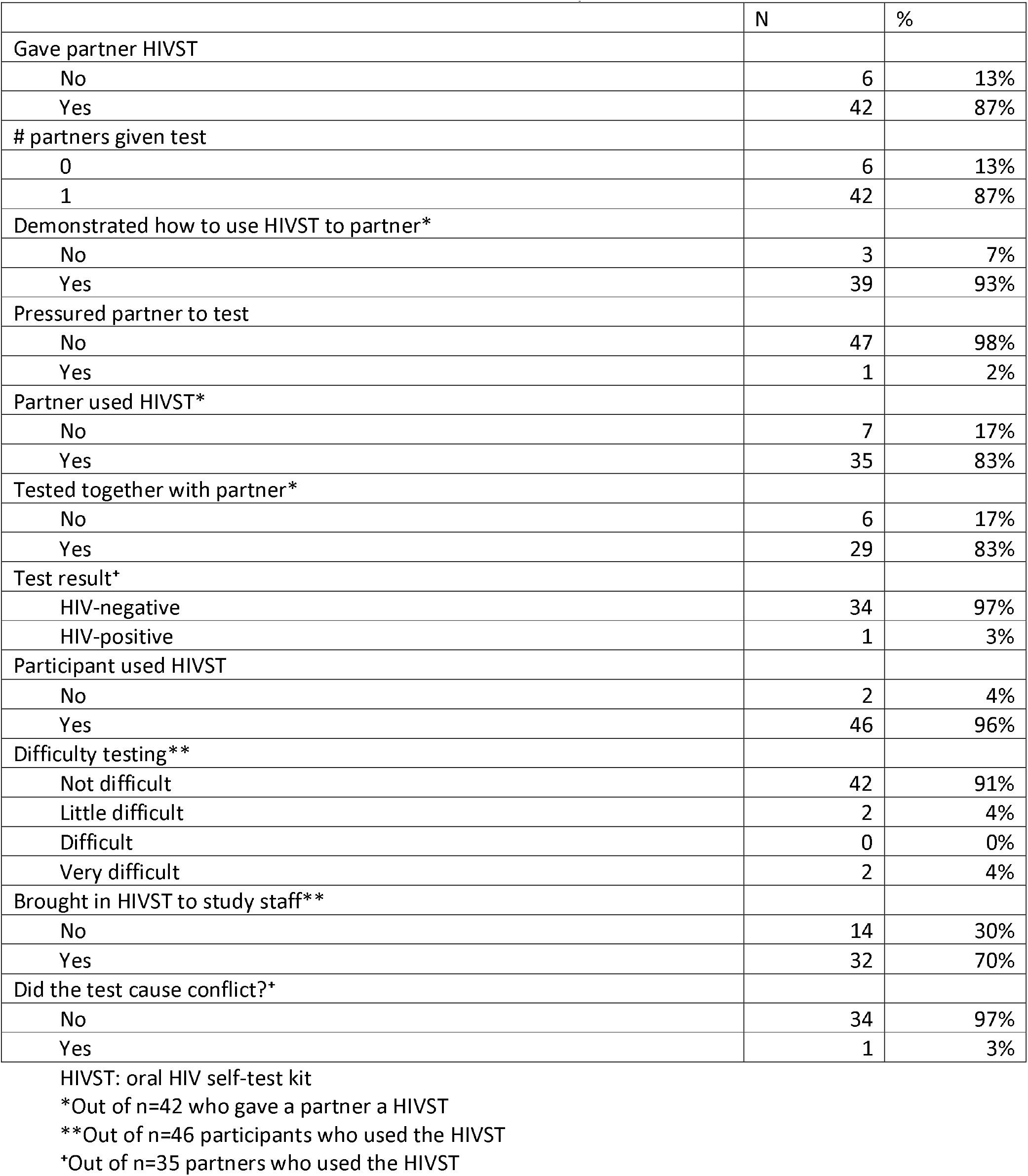
Results from HIV self-testing in women who returned for postpartum intervention arm in Cape Town, South Africa (n=48 women who returned for follow-up visit)

Women reported that the urine TFV test took 4-7 minutes for 96% of women. All women said that they understood the results, and 98% said they would like to get the test again in the future. However, 5 women stated that the result was not as they expected, all of whom had received a negative tenofovir urine test result, indicative of missing PrEP in the past 48-72 hours. Among participants without tenofovir detected, 64% were very satisfied or satisfied (**Table 3**).

**Table 3.**
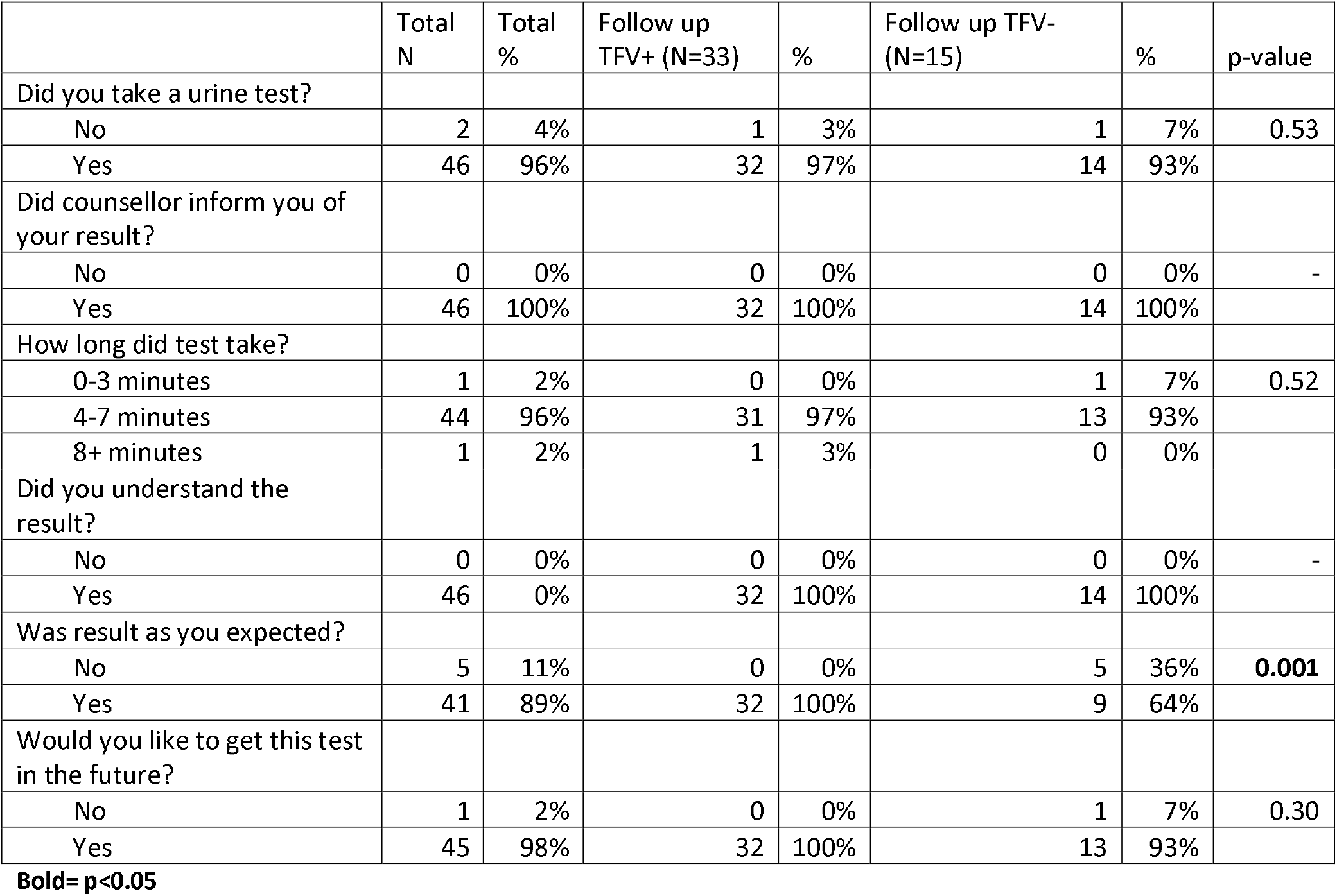
Acceptability of urine tenofovir testing in postpartum women on PrEP (n=48) in Cape Town, South Africa.

## Discussion

This is one of the first studies to evaluate the impact on PrEP adherence and continuation following HIVST and adherence biofeedback in postpartum women using PrEP. Our pilot trial of a combination intervention (HIVST for PrEP users and their partners plus biofeedback counselling) almost doubled recent PrEP adherence in postpartum women on PrEP (62% vs. 34%). Partner testing almost quadrupled in the intervention versus SOC arm (66% vs. 17%). The majority of women in the intervention arm tested together with their partner, giving them an opportunity to discuss their HIV status together as a couple. Postpartum women in the intervention arm reported high levels of satisfaction with both biofeedback counselling and HIVST.

These findings are important for the field of PrEP among postpartum and breastfeeding women who may be more at risk of poor PrEP adherence.^12^ In our parent study, PrEP-PP, there are noticeable drop-offs in clinic visits, PrEP collection, and adherence in postpartum women, mostly because they are no longer attending the clinic for ANC and most women do not know their partner’s serostatus and have limited partner communication about HIV and PrEP.^17^ The HIVST and biofeedback counselling used in our combined intervention may address these barriers through several complementary mechanisms, although further research is needed to fully understand *how* and *why* the combination intervention was effective in this pilot. First, HIVST for PrEP users may facilitate women monitoring her own status and therefore increase agency and ownership over her own care,^30^ while simultaneously reducing PrEP clinical visit time. At the same time, urine assays used for immediate biofeedback counselling may confront consequences of poor adherence. Women may not really believe missing doses would impact any impact. The biomarker may confront them with the consequence of not taking the medication as having no drug in their system. When done with a client-centered approach, we hypothesize that biofeedback counselling may improve counselling on barriers to daily PrEP use, resulting in greater patient understanding about adherence and strategies to address barriers to care, as well as improve patient-provider relationships as both work together to improve PrEP adherence and continuation. For adherent clients, visual positive feedback may promote sustained adherence, especially when coupled with a negative HIVST result. In other recent trials that provided drug feedback to PrEP users, acceptability was high. ^31-33^ Finally, HIVST for sex partners increases knowledge of partner status and therefore risk of HIV acquisition, and may promote increased communication about HIV and PrEP within couples, facilitating partners who may not attend health facilities to engage with HIV services in postpartum.

Importantly, the urine tenofovir testing in our intervention was provided in a context of ongoing counselling, and supportive non-judgemental approaches for PrEP clients. Counsellors were trained to interpret the urine test results with their participants so that they could review the results, and the implications together. If the test was negative, indicating that the participant missed PrEP dosing in the past 48-72 hours, the counsellor worked with clients to develop alternative prevention plans, including the use of condoms consistently until the client has taken PrEP again for 7-days, in addition to developing strategies to adhere to daily oral PrEP in the future. This technique was meant to remove judgement of the participant and moved the conversation into troubleshooting how best to improve daily adherence, including daily reminders, PrEP disclosure to family members, and the importance of condom use if PrEP was not taken daily. Biofeedback counselling was offered in the context of a postnatal care program that was focused on HIV prevention in PBFW who are at higher risk of HIV acquisition.

In parallel, partner disclosure, encouragement to take PrEP due to partner disclosure and HIVST has improved PrEP adherence in prior studies.^12,34,35^ Secondary HIVST distribution to sex partners may improve disclosure of PrEP use and adherence. Our study did not find any reports of intimate partner violence following testing. One woman reported “conflict” following testing with her partner who did not initially want to test. This result is in line with prior studies that show minimal adverse events, including intimate partner violence, were associated with HIVST distribution by female partners.^36-39^ Of note, the two women in our study who had partners diagnosed with HIV did not report IPV or any conflict following testing.

Findings are similar to recent work focused on use of retrospective feedback (not point-of-care) pharmacokinetic results to improve adherence counselling among non-PBFW using PrEP. In the 3P study in Cape Town young women, participants received retrospective feedback about PrEP drug levels at months 1-3 and showed high levels of adherence; yet only 8% and 5% in the incentive and SOC groups had detectable drug levels at 12-months.^40^ Women in the VOICE trial in southern Africa were interviewed post study and shown their PrEP adherence patterns. This disclosure showed that providing objective results stimulated discussions regarding adherence and women suggested that real time drug monitoring could improve adherence. ^19,20^ Prior studies have demonstrated the importance of adherence counselling on PrEP and simplified PrEP collection for optimal PrEP use.^23,41,42^

Discrepancies between self-report and biomarker results for recent adherence were significantly lower among women in the intervention arm compared to SOC arm (17% versus 46%), suggesting that biomedical measures of adherence encouraged honesty and transparency between patient and provider regarding adherence concerns. Importantly, providers were trained to communicate biofeedback results in a non-judgemental manner, and to use client-centred approaches to adherence education when there is discrepancy between self-report and biomarker adherence results. Almost all women in the intervention arm reported that the biofeedback result was as they expected (89%) and desired to keep receiving biofeedback counselling (including the urine assay test). However, women with discrepant results were less satisfied with the intervention and more likely to be non-adherent (did not have tenofovir in their urine). Overall, satisfaction with the combination intervention in women in the study was high.

Limitations of this study include the preliminary nature and modest sample size fundamental to a pilot study. The study was within another cohort study so is not integrated into standard postnatal or PrEP services. The differential loss to follow up may affect the acceptability of the intervention as we do not know why the women in the intervention group did not return, and we inferred that they were not adherent to PrEP to mitigate the potential bias in follow-up. Further, we are unable to determine which intervention, HIVST, biofeedback or both, impacted on women’s adherence, a common concern in combination interventions. The study was unblinded so participants may improve their adherence because of social desirability.

## Conclusions

We found that a combination intervention with HIVST for PrEP users and their partners plus biofeedback counselling using urine tenofovir testing doubled postpartum women’s recent adherence to PrEP. Partner HIV testing increased by four-fold in the intervention arm and discrepancies between self-report and biomedical measures of adherence were significantly lower in the intervention arm. These preliminary results suggest that simple combination interventions may be highly effective in improving PrEP adherence in this population, and more research is needed to understand the mechanisms of action, scalability, and longer-term effect of the intervention.

## Data Availability

Data available upon request to study PI

## Conflict of interest

None declared

## Acknowledgments

We would like to thank our study participants, PrEP-PP study staff and the City of Cape Town Department of Health staff.

## Funding

This study was supported through grants from the National Institute of Mental Health (TC and LM; R01MH116771) and Fogarty International Center (DJD; K01TW011187). We received study drug (Truvada®) from Gilead Sciences (CA, USA) and GeneXpert® test kits for STIs from Cepheid Inc. (CA, USA).

## Authors’ contributions

DJD: Designed the study, analysed data, wrote manuscript, approved final submission

KD: Designed the study, reviewed manuscript and approved final submission

RM: Cleaned and analyzed data, reviewed manuscript and approved final submission

DN: Analyzed data, reviewed manuscript and approved final submission

NM: Oversaw study implementation, reviewed manuscript and approved final submission

LGB Designed the study, reviewed manuscript and approved final submission

PG Designed the study, reviewed manuscript and approved final submission

TC: Designed the study, reviewed manuscript and approved final submission

LM: Designed the study, reviewed data and manuscript drafts, approved final submission

## Notes

### Competing Interest Statement

The authors have declared no competing interest.

### Clinical Trial

NCT04897737

